# Process Evaluation of APPLE-Tree (Active Prevention in People at risk of dementia through Lifestyle bEhaviour Change and Technology to build REsiliEnce), a Dementia-Prevention Study focused on Health and Lifestyle changes

**DOI:** 10.1101/2025.05.16.25327783

**Authors:** Elenyd Whitfield, Claudia Cooper, Harriet Demnitz-King, Sedigheh Zabihi, Julie Barber, Mariam Adeleke, Rachel M Morse, Amaani Ahmed, Alexandra Burton, Iain Lang, Penny Rapaport, Anna Betz, Zuzana Walker, Jonathan Huntley, Helen C Kales, Henry Brodaty, Karen Ritchie, Elisa Aguirre, Michaela Poppe, Sarah Morgan-Trimmer

## Abstract

**Introduction:** In a mixed-methods process evaluation embedded within a randomised controlled trial, we aimed to investigate how the APPLE-Tree secondary dementia prevention intervention might support behavioural and lifestyle goal attainment, through determining contexts influencing engagement and testing intervention theoretical assumptions.

**Methods:** We measured intervention reach and dose. We selected interviewees for setting (urban/rural, NHS/non-NHS), gender and ethnic diversity, from the 374 APPLE-Tree trial participants randomised to the intervention-arm. We interviewed 25 intervention participants (including six who withdrew), 12 facilitators and three study partners (family members or friends). Additionally, we analysed 11 interviews previously conducted during or after intervention delivery for an ethnography and 208 (55.6%) facilitator-completed participant goal records. We thematically analysed data, combining inductive/deductive approaches informed by the COM-B (Capability, Opportunity, and Motivation) behaviour change model. We audio-recorded a randomly-selected tenth of sessions and rated fidelity.

**Results:** 346/374 (92.5%) intervention-arm participants received some intervention (reach), and 305/374 (81.6%) attended ≥5 main sessions (predefined as adhering: dose). According to facilitator records, participants met a mean of 5.1/7.5 (68.3%) goals set. We generated three themes: (1) Building Capability and Motivation: Increased capability and reflective motivation altered automatic motivation; (2) Connecting with other participants and facilitators helped increase social opportunity, motivation and capability; (3) A flexible, tailored approach increased capability, motivation, and opportunity for engagement.

**Conclusion:** The intervention supported behaviour change, through increasing knowledge and providing space to plan, implement and evaluate new strategies and make social connections. Feedback indicated that the intervention was flexible and inclusive of diverse preferences and needs.

## Introduction

The number of people with dementia is expected to increase to 153 million worldwide by 2050 (1). Many dementia risk factors are potentially modifiable, including cardiometabolic factors, physical inactivity and social isolation (2). Nearly half of individuals who consult primary care with memory loss symptoms develop dementia within three years (3). While many receive lifestyle advice around exercise, diet, social and cognitive stimulation, there is limited evidence to support specific interventions (4,5). Compelling evidence for the impact of lifestyle or psychosocial interventions on cognitive decline in people at increased dementia risk comes from a Randomised Controlled Trial (RCT) involving a 2-year intensive intervention, delivered by experts, promoting nutrition, exercise, social connections, cognitive training and vascular risk management (6).

It is estimated that delaying Alzheimer’s disease onset by a year will reduce the number of cases in 2050 by 11% worldwide (7), so the potential benefit of secondary dementia prevention is considerable. To make an important difference, such interventions need to be widely acceptable; realistic in terms of the resources required to be deliverable at scale; and inclusive of those experiencing socioeconomic deprivation and other, including ethnic, minoritised groups who are at increased dementia risk (8,9).

The APPLE-Tree dementia prevention intervention was co-designed to be acceptable, inclusive and delivered at scale. The APPLE-Tree RCT recruited participants between 2020 and 2022; its logic model (10) and pilot study (11) are reported. In this process evaluation, conducted before knowing trial results, we aimed to investigate (i) intervention reach, dose and fidelity, (ii) contexts influencing engagement, and (iii) alignment of findings with theoretical assumptions about how the intervention might have supported participants to meet personalised behavioural and lifestyle goals.

## Methods

### Study design

We used the Medical Research Council (MRC) guidance on evaluating complex interventions (12). Based on the design and development of the logic model (10), we selected a theory-driven mixed-methods approach. London (Camden and Kings Cross) Research Ethics Committee (Reference: 19/LO/0260) and UK Health Research Authority approved this study in April 2019. The protocol is registered (ISRCTN17325135). We designed the sampling strategy and topic guides to test *a priori* theories about how the intervention works and causal assumptions regarding intervention mechanisms developed during co-design and iterated by the research and Patient and Public Involvement (PPI) groups.

### Setting and sample

The APPLE-Tree Trial recruited 748 people aged 60+ with subjective or objective cognitive concerns, without dementia, by mail outs from participating general practices, memory clinics and social and print media (10). Participants gave written or audio-recorded informed consent for the process evaluation, prior to randomisation using 1:1 allocation to the intervention or control arm (provision of dementia prevention information). As one participant was randomised twice in error and subsequently withdrawn after consulting the Clinical Trials Unit and independent statistician, the sample size was 746, of whom 374 were randomised to the intervention arm.

APPLE-Tree was delivered to groups of between four and nine participants. We initially approached all participants and facilitators from four intervention groups, purposively selected to encompass urban and rural, NHS and non-NHS contexts. Study partners (family members or friends who supported participants’ involvement) were also invited to participate in an interview with a researcher. We recruited additional participants from minority ethnic groups, and female participants, to increase sample diversity. Participants who withdrew from the intervention, but who agreed to continue completing research outcomes were also invited to interview. Sample size for the interviews was planned (estimated *a priori* as 45 total) to ensure sufficient diversity for setting (urban/rural, NHS/non-NHS), gender and ethnic diversity (18).

In addition to the primary interviews, secondary analysis was carried out on eleven interviews conducted with APPLE-Tree intervention arm participants during and after the intervention in an embedded visual ethnography study (19).

### Intervention description

The APPLE-Tree intervention aimed to promote healthy lifestyle, increase pleasurable activities and social connections and improve long-term condition self-management. Facilitators delivered ten, one-hour group video call sessions over six months (fortnightly), alternating with ten video-call ‘tea breaks’ (less structured, facilitated social sessions); facilitators conducted individual goal-setting phone calls fortnightly, to a SMART (Specific, Measurable, Achievable, Relevant, and Time-Bound) guide. Full adherence was defined *a priori* as receiving at least five main sessions. From months six to 12, participants met monthly to discuss how they were implementing learnt strategies, with those not attending receiving monthly goal-setting phone calls, or, if they preferred discussion by email. Participants had access to a website that included cognitive training and received a single food delivery and a pedometer.

The intervention was delivered by 16 university-employed facilitators, psychology or social science graduates, paired with 15 third sector or NHS-employed facilitators; or at two sites, by two NHS-employed facilitators. All facilitators (31 in total) were non-clinically trained. They attended training and group supervision fortnightly with a clinical psychologist (SB) and monthly with a nutritionist (ABe). Further intervention details have been published elsewhere (10). A TiDiER (Template for Intervention Description and Replication) is provided in Appendix A.

### Data collection

*Qualitative interviews* were conducted after completion of the main intervention groups (except two participants interviewed after sessions six and seven respectively) by researchers not involved in intervention delivery or outcome assessment. Interviewees who withdrew from the intervention were interviewed in the month after making this decision. Topic guides (see Appendix B) drew on the intervention logic model (Appendix C) (10) to explore how participants experienced the intervention, how it might have supported them to make and maintain lifestyle changes, and perceived impact of these changes.

#### Additional data analysed

EW reviewed transcripts of 11 participant interviews from a visual ethnography conducted during the intervention (March-September 2022) to identify data potentially relevant to our aims, which were included in this analysis (14).

Facilitators recorded goals set, their content and whether they were achieved using a standardised spreadsheet, on which they made written notes during or immediately after goal calls. We randomly selected (using random number generation) a group intervention session to audio-record from each intervention group (n=41) (generating a random sample of one tenth of all intervention sessions delivered, as there were ten main sessions) to assess facilitator adherence to procedures, using a standard fidelity checklist approach similar to one developed for a previous intervention (15). Information on participants’ use of cognitive training was downloaded from the study website.

## Analysis

We compared the trial and process evaluation samples, to each other and the 2021 census data for England and Wales (16) and the English Housing survey (17). We described intervention reach, adherence and fidelity using summary statistics. We analysed goal content at participant level, reporting numbers of goals that corresponded with goal themes, generated through content analysis. We thematically analysed the interviews. Interviews were coded in NVivo by EW, deductively and inductively, drawing on COM-B model concepts (16). In this model each modifiable factor (Capability, Opportunity, Motivation leading to a Behaviour) is comprised of two parts. Psychological capability includes knowledge and understanding; physical capability refers to physical attributes and bodily capacities; reflective motivation to conscious processes such as evaluation and planning; automatic motivation to desires, habits and impulses; and opportunity to the physical and social contexts of behaviours (16). The codebook and three de-identified transcripts were discussed with co-authors CC, SM-T, RMM and AA.

SB listened to the audio recordings of intervention sessions, completing fidelity checklists. We calculated the proportion of expected intervention components delivered using the fidelity checklist. We rated fidelity according to established thresholds, with 81–100% constituting high fidelity. On a 5-point scale (1– not at all to 5-very much) we rated whether facilitators kept the group focused, participants engaged in each intervention component, and the session kept to time.

The authors assert that all procedures contributing to this work comply with the ethical standards of the relevant national and institutional committees on human experimentation and with the Helsinki Declaration of 1975, as revised in 2013.

## Results

### Quantitative findings

#### Reach and dose

Compared to Census data for people aged 65 years and above, trial and process evaluation samples included more people from non-White ethnic groups (11.6% and 13.9% respectively, relative to 6.4% in the Census); and living as a couple (66.5% and 63.9% versus 60.7% in the Census)(16). While 80% of people in England aged 65+ are home owner occupiers, this was higher in the trial (88.0%) and process evaluation (83.3%) samples (17).

346/374 (92.5%) intervention arm participants received one or more main intervention sessions; 305/374 (81.6%) of participants randomised to the intervention arm attended five or more main sessions, and 89/374 (23.8%) attended all 10 main sessions. 2363/3279 (72.1%) possible tea breaks and 983/1903 (51.7%) of the monthly catch-up groups, scheduled between 6-12 months occurred. 57/374 (15.2%) participants withdrew from the intervention. 49 (13.1%) participants randomised to the intervention used the cognitive training app. 13 (3.5%) participants accessed the intervention using a device provided by the study team; all others used their own devices.

#### Fidelity

Mean rater fidelity scores out of five were: 4.69 for ‘Keeping the group focused on the manual, 4.51 for ‘Keeping participants engaged’ and 4.97 for ‘Keeping the session to time’. Overall, fidelity was high (94.5%; 14.17/15).

#### Goal data

We had useable goal-setting call data for 208/374 (55.4%) participants, who achieved an average of 7.4 goal calls and set on average 7.5 goals, with a range of 0-23 goals set. On average, 5.1 (68.3%) goals were recorded as met in the first six months of the intervention. A small number of goal calls also acted as catch-up sessions. The most common reason for missing goal data was that external (third sector) facilitators did not provide it to the study team. Participants made goals related to: improving diet (n=203); increasing exercise (n=189); relaxation, mood and wellbeing (n=90); planning activities (n=79); improving physical health through other strategies (n=77); sleeping better (n=63); engaging with pleasurable activities (n=64), including creative and artistic activities (n=24); cognitive stimulation (n=51); connecting with others (n=47); engaging with the programme; reducing alcohol (n=24) or smoking (n=2); completing other tasks (n=18); managing memory issues (n=11); and reducing screen time (n=6).

### Interview sample description

EW (social science researcher), research assistants who facilitated the intervention with participants in other groups, and SB (clinical psychologist) conducted interviews between September 2021 and May 2023 with 40 participants: 19 participants who completed and 5 who did not complete the intervention (one withdrawer returned), goal data were available for 10/19 and 3/5 respectively; 12 facilitators and 3 study partners: two daughters and one wife of a participant, all White UK ethnicity. Additionally, we included 11 interviews from a previous visual ethnography in the analysis (14).

Table 1 shows that relative to the baseline trial population, those interviewed for the process evaluation were more likely to be female and from a non-White UK ethnic group. Table 2 summarises process evaluation data.

**Table 1:** Characteristics of the interview sample compared to the baseline Trial population^1^.

|  | All (n=746) | Interview sample (n=36) |
| --- | --- | --- |
| Age – mean (SD)<br>range | 74.4 (6.9) (**n=742**)<br>60.1 to 102.7 | 74.6 (6.3) (**n=36**)<br>61.6 to 90.6 |
| Sex | **743** |  |
| Male | 392 (52.8%) | **36** |
| Female | 350 (47.1%) | 16 (44.4%) |
| Other | 1 (0.1%) | 20 (55.6%) |
| Ethnicity | **742** | **36** |
| White UK | 599 (80.7%) | 28 (77.8%) |
| White other | 57 (7.7%) | 3 (8.3%) |
| Asian | 50 (6.7%) | 2 (5.6%) |
| Black | 13 (1.8%) | 2 (5.6%) |
| Mixed | 16 (2.2%) | 1 (2.8%) |
| Arab | 3 (0.4%) | 0 (0.0%) |
| Other | 4 (0.5%) | 0 (0.0%) |
| Marital Status | **742** | **36** |
| Single | 46 (6.2%) | 1 (2.8%) |
| Married/civil partnership | 464 (62.5%) | 21 (58.3%) |
| Living with partner | 30 (4.0%) | 2 (5.6%) |
| Widowed | 108 (14.6%) | 8 (22.2%) |
| Divorced | 92 (12.4%) | 3 (8.3%) |
| Unable to specify | 2 (0.3%) | 1 (2.8%) |
| Highest level of education | **742** | **36** |
| No Education | 2 (0.3%) |  |
| Primary | 10 (1.3%) | 1 (2.8%) |
| Secondary (e.g. O level;<br>GCSE) | 167 (22.5%) | 11 (30.6%) |
| Further (e.g. A level; BTEC;<br>NVQ) | 199 (26.8%) | 7 (19.4%) |
| Degree | 204 (27.5%) | 7 (19.4%) |
| Postgraduate | 147 (19.8%) | 10 (27.8%) |
| Other | 11 (1.5%) | 0 (0.0%) |
| Unable to specify | 2 (0.3%) | 0 (0.0%) |
| Employment | **742** | **36** |
| Full time employment | 32 (4.3%) | 2 (5.6%) |
| Part time employment | 54 (7.3%) | 1 (2.8%) |
| Retired | 607 (81.8%) | 31 (86.1%) |
| Unemployed/unable to<br>work | 17 (2.3%) | 0 (0.0%) |
| Other | 29 (3.9%) | 2 (5.6%) |
| Unable to specify | 3 (0.4%) | 0 (0.0%) |
| Living situation | **742** | **36** |
| Live alone | 198 (26.7%) | 10 (27.8%) |
| Live with other relatives | 532 (71.7%) | 26 (72.2%) |
| Live with friends/other<br>people | 6 (0.8%) | 0 (0.0%) |
| Other | 6 (0.8%) | 0 (0.0%) |
| Type of accommodation | **742** | **36** |
| Council Rented | 33 (4.4%) | 2 (5.6%) |
| Private rented | 40 (5.4%) | 2 (5.6%) |
| Own home | 653 (88.0%) | 30 (83.3%) |
| Supported living | 12 (1.6%) | 2 (5.6%) |
| Other | 4 (0.5%) | 0 (0.0%) |
| Diagnosis | **742** | **36** |
| Mild Cognitive Impairment | 125 (16.8%) | 8 (22.2%) |
| Memory Concerns or problems | 59 (8.0%) | 2 (5.6%) |
| Other | 13 (1.8%) | 0 (0.0%) |
| Not given diagnosis | 525 (70.8%) | 25 (69.4%) |
| Unable to specify | 20 (2.7%) | 1 (2.8%) |

**Table 2:** Description of process evaluation data by group.

|  |  |  |  |  |  | ***Additional***<br>***primary***<br>***interviews*** | ***Secondary***<br>***interviews***<br>***analysed*** |
| --- | --- | --- | --- | --- | --- | --- | --- |
|  | Group 1<br>(NHS,<br>suburban)<br>N=3 | Group 2<br>(non-NHS,<br>rural)<br>N=3 | Group 3<br>(non-NHS,<br>urban/<br>suburban)<br>N=3 | Group 4<br>(non-NHS,<br>urban/<br>suburban)<br>N=3 | Participants<br>(withdraw)<br>N=6 | Group 5<br>(Recruited<br>from<br>multiple<br>interventio<br>n groups)<br>N=6 | N=11 |
| **Participants**<br>**(completed**<br>**intervention)** | 3 | 3 | 3 | 3 | 1<br>(1 rejoined<br>following the<br>interview) | 7 | 11 |
| **Goal data available** | 3 | 2 | 2 | 0 | 3 | 3 | 7 |
| Gender | 75% male<br>25% female<br>0% other | 100% male<br>0% female<br>0% other | 100% male<br>0% female<br>0% other | 33.3% male<br>66.6% female<br>0% other | 16.6% male<br>83.3%female<br>0% other | 42.8% male<br>57.1%<br>female<br>0% other | 72.7%<br>female<br>27.2% male<br>0% other |
| Ethnicity: White UK | 100% | 100% | 100% | 66.6% | 100% | 71.4% | 54.5% |
| White other | 0% | 0% | 0% | 0% | 0% | 14.2% | 18.1% |
| Asian | 0% | 0% | 0% | 0% | 0% | 14.2% | 9% |
| Black | 0% | 0% | 0% | 33.3% | 0% | 0% | 9% |
| Mixed/other | 0% | 0% | 0% | 0% | 0% | 0% | 9% |

### Qualitative findings

#### Theme 1: Building Capability and Motivation: Increased capability and reflective motivation altered automatic motivation

The intervention increased capability through supporting learning, and reflective motivation by facilitating evaluation and planning. Learning was primarily through main group sessions and an accompanying course book. Evaluation and planning were facilitated through goal setting. Discussions of ideas, plans and achievements took place in group sessions and tea breaks; with ‘modelling’ behaviours as a ‘motivational device’(18). In subthemes below, we describe how the intervention increased (1) capability, (2) reflective motivation and (3) automatic motivation.

##### Subtheme 1: Increased capability

Participants learned new information about healthy behaviours and the science of familiar healthy messages. Facilitators and participants described how the intervention ‘reinforced’ familiar positive health messages. Some participants, including a participant who withdrew felt information was insufficiently novel; a facilitator describes how they responded to this:

> “Well, I already know that” and that’s if you leave it at the content level as you already know. If you take it to a meta level, “Yes, we already know that” but it’s “This is why it’s relevant when seen through the APPLE-Tree lens, why it’s important, and thinking about MCI [Mild Cognitive Impairment], brain health” (facilitator, NHS Trust).

Goal calls were an opportunity to explore barriers to healthy habits:

> It was helpful to then use the goal calls to work through what barriers, why aren’t they doing it, why would they do it? And do it that way (facilitator, research assistant).

The study partner of a participant felt their father was more open to new ideas now, and described him taking greater responsibility for his own health and wellbeing, suggesting an increased agency and capability:

> “he is quite happy to go to the supermarket now. Something as basic as that, whereas before my mum would do it, but he’ll quite happily go himself now, if that makes sense. And he’ll go to shops and do what he needs to do. He’ll make his own doctor’s appointments rather than my mum doing it. So, there is a few things that he does do himself now which he didn’t do six months ago.” (study partner of father in intervention, 60s).

Among people who withdrew from the intervention, disjuncture between expectations and course content was a common theme. They expressed surprise there was not more focus on cognition and memory, which was also felt by some participants who completed the course. One participant was motivated to make lifestyle changes but felt that her physical health problems and living alone prevented engagement with the strategies suggested.

##### Subtheme 2: Reflective motivation

New information was described as “a way of giving that control”, prompting reflective motivation around health improvements; “how do I look at what I do and decide is that good for me” (facilitator, NHS Trust).

A facilitator described the ‘little epiphanies’ that were happening in their group/s when people began implementing healthy behaviours. For some, there was a sense of empowerment by the group sessions and other intervention elements. For example, one participant reported increased confidence to engage in activities; she described how the intervention supported her to reengage after the pandemic lockdown:

> “I wasn’t going anywhere, I wasn’t travelling to London, I wasn’t going in the car, I was doing nothing. But slowly, together with the COVID changing, and this research giving me confidence, that I’ve come out of it. I’ve been to two or three exhibitions now.” (participant: secondary analysis, female, age unknown).

One participant wore her pedometer in bed to include steps when she gets up in the night saying it is “part of me now” (participant: secondary analysis, female, aged 60-70). Increasing step count was a popular goal. An alcohol unit calculator included in one intervention session was also described as constructive by a facilitator, allowing people to take “more power back over better decisions and their choices” (facilitator, research assistant). Few intervention arm participants (n=49) used the cognitive training app, many citing technical barriers.

##### Subtheme 3: Automatic motivation

These increases in capability and reflective motivation enabled changes in habits (automatic motivation). This included eating more nuts, less sugar, more olive oil, adopting a “brain healthy” diet, taking supplements, reading food labels, reducing alcohol, increasing exercise, drinking more water, checking blood pressure, and joining new social activities. Some participants reported health benefits that in turn increased capability, such as weight loss, more energy, stamina, and improved sleep; one participant reported reversing their pre-diabetes. These changes align with our *a priori* logic model.

A participant spoke of how the programme helped her to develop better habits:

> Being aware of it and putting it into your daily routine are two different things, and this study has made, how shall I put it, made me take responsibility … I have started introducing the changes in my diet and lifestyle … this is helping because every week having this check in, the tea breaks and all the things, so it is kind of helping me to form that habit (participant: group 5, female, aged 70-80).

Some participants were encouraged to return to activities that they had enjoyed but discontinued, including dancing, tai chi, jogging and swimming:

> It’s made me realise that dance has always been a big thing for me and I don’t play enough music and I don’t jig about enough as my ability would allow… But I only have to hear music and I want to jig about to it. And that’s good for the soul as well as the body (participant: group 5, female, aged 80-90).

A participant who had had a hip replacement and knee operation was supported to recommence jogging in his late 70s. One participant faced her longstanding fear of swimming following a traumatic incident as a child, which she described as “a very big step” (participant: group 5, female, aged 70-80).

Some participants also described changes in outlook, increased confidence and self-efficacy and empowerment, reflecting increased psychological capability. In this next example, a participant described reflecting more on how his daily activities affected him:

> How did my mood or my mentality change because of the adaptation of what happened? So it’s a lot of things. Instead of looking at things blinkered, I now look at things in a very wide sphere (participant: group 2, male, aged 60-70)

A facilitator described participants who had caring responsibilities that left little time for themselves. She felt that the intervention had been “fantastic for them in reclaiming back their own autonomy” (facilitator, third sector).

The role played by participants’ social context in dietary changes was variable. Some partners actively supported positive dietary changes; one felt she had “sort of been in partnership” with the programme and was very pleased at the changes her partner made (study partner, group 2 participant’s wife, husband aged 70-80). However, habits and tastes of partners could also be a barrier to possible changes because shopping, cooking and eating are often shared, habitual activities.

#### Theme 2: Connecting with other participants and facilitators helped increase social opportunity, motivation and capability

The APPLE-Tree groups created connections with others, motivating people to engage with the intervention, and institute behaviour change, as exemplified here:

> One of the big positives for me that it was in a group environment rather than one on one. It’s nice to be able to share ideas and listen to other people’s experiences and to share your own ideas and experiences obviously. You don’t feel so isolated (participant: group 5, male, aged 60-70).

A participant described the importance of having shared lived experience with other participants and how the groups helped him realise that he was not “the only one” experiencing memory problems:

> We’re all in the same boat. But you don’t realise until you actually discuss it amongst yourselves, yeah. You just button it up (participant: group 1, male, aged 60-70).

A facilitator gave examples of participants who, though initially sceptical about the programme, valued the social aspect:

> Perhaps people who felt that their memory problems were quite isolating for them and felt quite a lot of frustration that perhaps their family didn’t understand what they were going through or felt embarrassed to talk about it, this was finally a space where they could talk openly about it, knowing that other people were experiencing similar things (facilitator, research assistant).

Facilitators and participants described how some groups/group members met up in person, or in one case, in their own support group online after the programme; a few members of another group continued to meet at a local café.

> It was quite a good mix of interests and abilities and I found I had things in common with several of them. And we are going to continue to meet together which is always a good thing. (participant: female, aged 80-90)

Positive relationships with group members motivated engagement. A participant commented:

> It’s something I look forward to because it’s a change of scenery, it’s a different type of dialogue with different friends, so that’s a bonus as well (participant: group 2, male, aged 60-70).

A facilitator expressed surprise at how important this social connection seemed to be:

> The social element was often one of the main facilitators of the group and it would often be the things that meant people would come back each week because they felt they were benefiting from being kept accountable each week to the things they want to try or the things they want to change or they’re getting to know other people (facilitator, research assistant).

Group discussions increased knowledge and motivation for behavioural change. For example, a facilitator recounted how one participants’ goal of decluttering their house inspired others. Participants influenced each other to do online tai chi, Pilates and yoga. A participant described how he found hearing other people’s coping strategies “really encouraging” (participant: group 3, male, aged 70-80). Another described how sharing ideas motivated him:

> Sharing those moments, sharing the different ideas, oh I didn’t think of that, that’s a good idea, I’m going to try that, is very, very useful (participant: group 5, male, aged 60-70).

A participant’s partner described conversations in groups as ‘invaluable’ by sharing “You could say wow, she’s doing that. What could I do” (study partner, group 2, wife to participant aged 70-80). This again speaks to the reflective motivation that can be prompted through discussion with others.

Facilitators described how some groups “gelled” more than others. A facilitator reflected on how, in her experience, this could inhibit levels of engagement:

> Because they hadn’t gelled so much, it just made the group discussions a bit wooden. So there wasn’t so much involvement and then they weren’t super personable and that meant that the goals and other things that were … surface-level, but it felt like they weren’t engaging as much as they usually would have (facilitator, third sector).

One participant felt that other group members were more physically capable than she was:

> “The exercise bits are very, very limited in what I can do. But there were little bits I could use and adapt. But they were the other end of the scale. You had people who were doing four hour fitness programmes either in the gym or walking for miles and miles every single day…. And one of the effects it had on me was at the end of a session, I felt I had failed. And I mentioned this to one of the leaders in the individual call.” (participant: group 5, female, aged 80-90).

A study partner described how their father enjoyed spending time with ‘completely different people from different walks of life’, outside of his usual ‘bubbles’. He has become more outgoing as a result: “He’s quite happy to sit with the other people who he doesn’t know, talk to people in the supermarket on the checkout and whatnot. So, he’s become a bit of a social butterfly” (study partner: group 1 participant’s daughter, father aged 60-70).

Group interactions in group 2 were reported positively by all participants and a study partner, who described how it had “brought [her father] out of his shell a bit more”, and he had “really enjoyed” the zoom meetings and tea breaks (study partner, participant’s daughter, father aged 80-90). Participants in this group met up in person.

Several participants who withdrew from the intervention because they did not find it helpful did not appear to have experienced this group cohesion; one stated that they were happy to talk one-to-one but not in a group. Most participants interviewed spoke very positively about their facilitators’ encouragement and approach. A participant who felt less comfortable speaking in the group sessions reflected on the ‘rapport’ they had built with the facilitator during goal calls: “I found I had no problem speaking quite openly. That I find a lot easier than doing it within the group” (participant: group 1, male, aged 70-80).

Good relationships enabled constructive goal setting, as described by a facilitator, who reflected on the challenge of balancing relationship building and goal setting in one-to-one calls. Participants who were living on their own often seemed to particularly value these relationships:

> There were some people at the table that live on their own in particular I ended up talking to a lot longer. They’d be 30 minutes, maybe even more, each time I had a goal call with them. But again, they were great, they set so many goals, it was perfect. But, again, I think there’s that fine line between having a chit-chat and setting goals. But it really helped to think about why they were doing it, and I think building a relationship made it better “(facilitator, research assistant).

Participants shared recipes, photographs and videos of their activities. Trying each other’s recipes built social connections. One participant, a yoga teacher, led a breathing exercise in a tea break, intending to repeat in online meetings after the group ended. Another participant led a group meditation.

#### Theme 3: A flexible, tailored approach increased capability, motivation, and opportunity for engagement

Facilitators were able to tailor the intervention to individual needs and preferences, which helped motivate participants. Participants valued the choice provided by the multiple intervention elements:

> The flexibility of what participants take from the intervention and what they leave, because they’re not going to take everything… one of the main things as a facilitator… is to find their aspects of the intervention that work for them … let them lead on what they want to make changes in, and what aspects work for them” (facilitator, research assistant).

Another facilitator reflected that there is ‘something for everyone’ in the course content, which helped to facilitate motivation for behavioural change:

> I think that’s what’s really good about APPLE-Tree is that in session one and two if there isn’t something for you there will be something for you in the later session to do with diet or exercise, that can kind of help with that. Or that will spark your interest and motivation, motivate you to make some changes (facilitator, research assistant).

One facilitator described responding to resistance to lifestyle change by finding something else a participant might value in the sessions, such as connecting with others, “finding something for everybody”. Another discussed ‘tweaking’ exercise videos to meet people’s needs, selecting lower intensity options for those with reduced mobility (facilitator, third sector).

Tea breaks were described as both “integral” to the intervention by a facilitator and a flexible space that differed between groups:

> Some groups really enjoyed the structure of recapping and talking about next week and … sharing recipe suggestions from other people in the groups. Then some of the tea breaks were people just really connecting with each other (facilitator, third sector).

They were an opportunity for participants to take more of an active role:

> I like the tea breaks because you could discuss things, whereas in the sessions, you were sort of taught, and in the tea breaks, you could talk about it (participant: additional, male, aged 70-80).

## Discussion

We describe how the APPLE-Tree intervention increased physical and psychological capability, through learning, with course content delivered via group sessions and the course book. Reflective motivation was increased through planning and evaluation. Goal calls were the primary space for this; discussions in group sessions were also important. The activity diary, included in the intervention booklet and discussed in main sessions, was used for planning and reflective evaluation. Changes in habits improved capability further due to physical health improvements. Social connections alleviated a sense of isolation around memory issues for some and motivated continued engagement through discussion and ‘modelling’.

Because groups were an agent of change, their composition mattered. This echoes and extends findings around the importance of social connection from the APPLE-Tree pilot study (11). By contrast, participants who did not find the intervention helpful did not experience groups as cohesive or helpful; some felt unconvinced of a need to change lifestyle, or that the intervention did not equip them with personally relevant strategies. Reflections from many participants however, and facilitators, indicated that the programme offered flexibility to personalise the intervention, enabling positive change through diverse pathways. Intervention dose, reach and fidelity were high.

The APPLE-Tree intervention aimed to reduce cognitive decline, with secondary outcomes to reduce anxiety, depression, and improve sleep, quality of life and functioning. Intended mechanisms include changes in diet, physical functioning, social networks and support (10). If forthcoming trial results indicated these outcomes were attained, insights from this process evaluation, and a recent pre-implementation study (19) will support larger scale implementation. If they do not, these findings may nonetheless inform secondary prevention. A previous intervention successfully addressed dementia risk factors (diet and reduced hypertension (20)) over two years and improved health-related quality of life but not the primary outcome of cognition, in older people recruited for increased vascular risk (21). Interventions that successfully support positive lifestyle changes can inform more effective health promotion, even if cognitive changes are not demonstrated.

*“The power of prevention*” is a focus of the Darzi independent investigation to inform the English NHS ten-year plan (22). Realising this ambition for secondary dementia prevention will require an acceptable, flexible intervention that supports personalisation. Delivery by non-clinical facilitators including social prescribers, band 4 NHS workers and third sector workers with skilled supervision (23) appeared to work well (24).

We captured diverse experiences, including accounts from six participants who withdrew, but there were some limitations. The intervention was only delivered in English within a well-resourced team. People living alone and in rented accommodation were under-represented in this trial, relative to the general population. In addition to structural barriers to research participation affecting people from socioeconomically deprived backgrounds in most trials (25), this may reflect their relative digital exclusion. All participants had memory concerns, so there may have been a recall bias towards more recent events. Use of contemporary goal call data partially mitigated this, though data were unavailable for 40% of participants. This could introduce bias, if external facilitators systematically differed in their goal-setting approaches to internal facilitators, though all received the same training. The trial took place during the pandemic and post-pandemic period; this context may have influenced how participants were able to use the intervention (26).

### Conclusion

The intervention was effective in facilitating capability, aided by psychoeducation and consequent learning and reflective motivation, promoting behaviour change and new habit formation. The group aspect and the promotion of group cohesion were important to this. Flexibility in the wide range of topics available, goal setting approaches, and the multiple components of the intervention enhanced its adaptability and potential usefulness.

## Trial registration

ISRCTN17325135. Registration date 27 November 2019.

## Authors

CC was chief investigator and JAB, AB, IL, PR,ZW, JH, HCK, HB, KR, EA, SMT co-investigators on the APPLE-Tree grant and involved in designing the study and formulating the research questions. EW, HDK, SZ, RMM, AA, AB and MP subsequently joined the team and supported the development of research questions for this process evaluation. MA was the junior trial statistician who supported quantitative aspects. All authors were involved in carrying it out, analysing and interpreting the data and writing the article, of which EW prepared the first draft.

## Funding

This research was carried out with funding from the Economic and Social Research Council (ESRC) and the National Institute for Health and Care Research (NIHR): ES/S010408/1.

## Declaration of Interest

None

## Data Availability

The data that support the findings of this study are available on reasonable request from the corresponding author, [EW]. The data are not publicly available due to their containing information that could compromise the privacy of research participants.

## Appendix 1

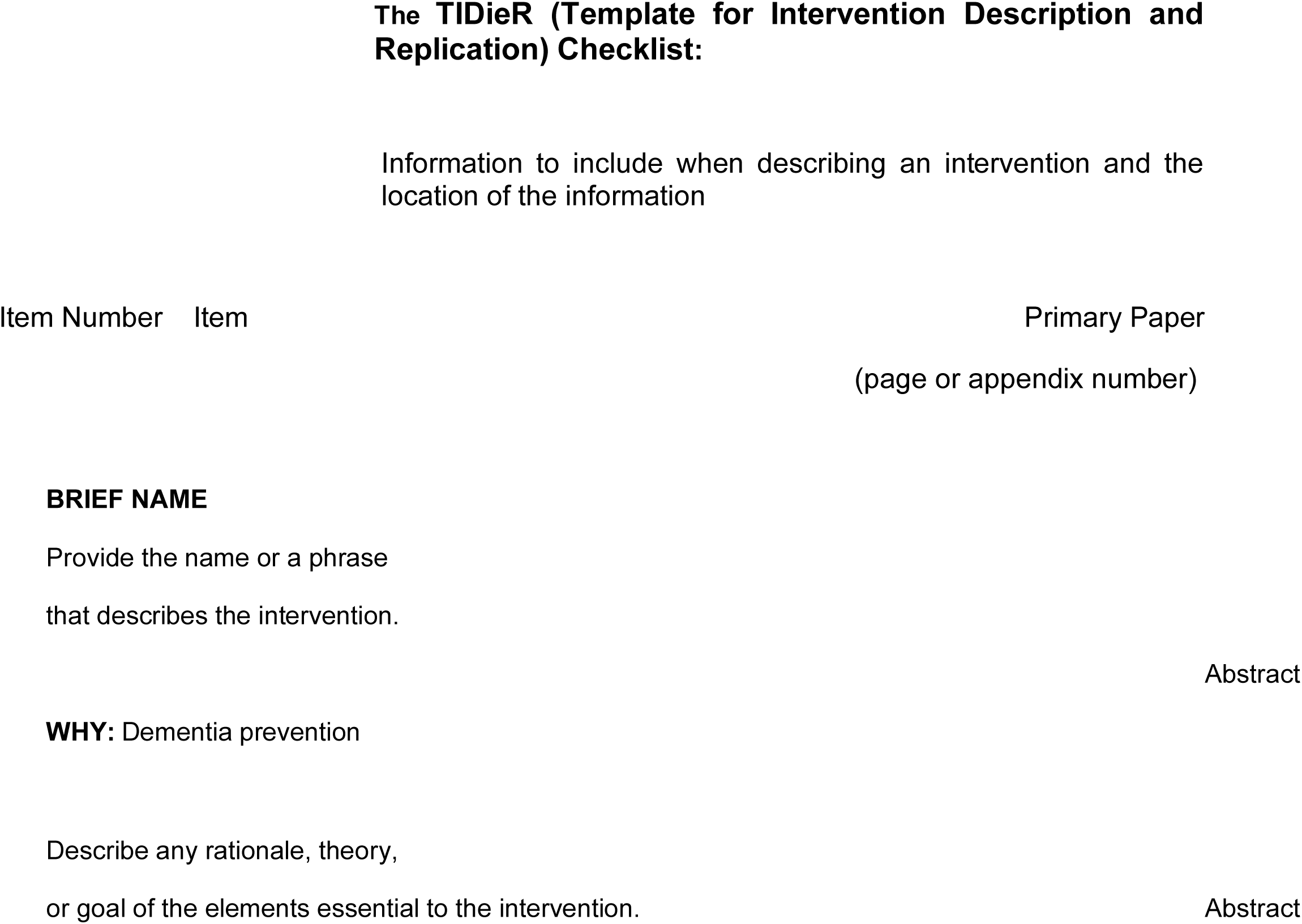
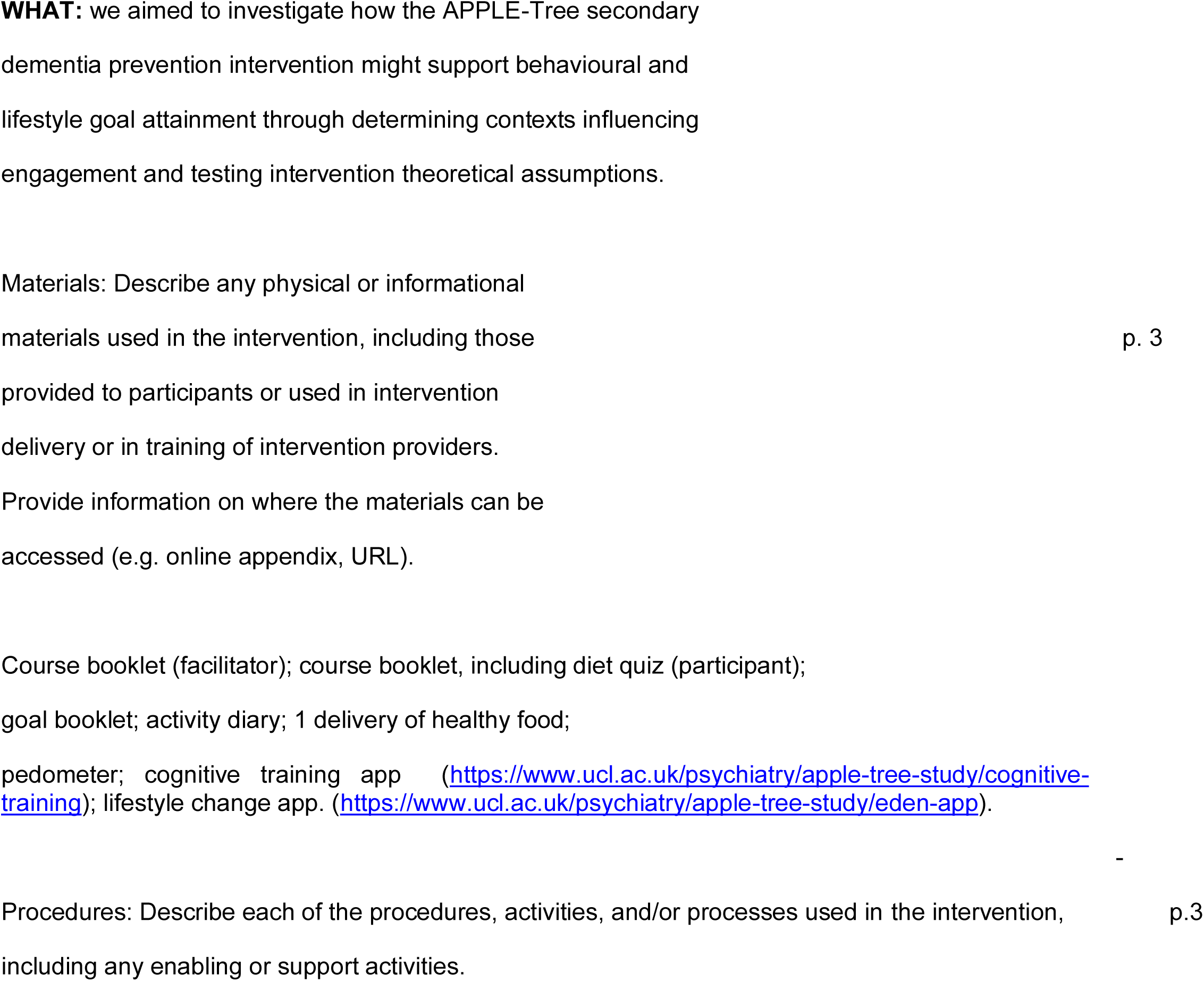
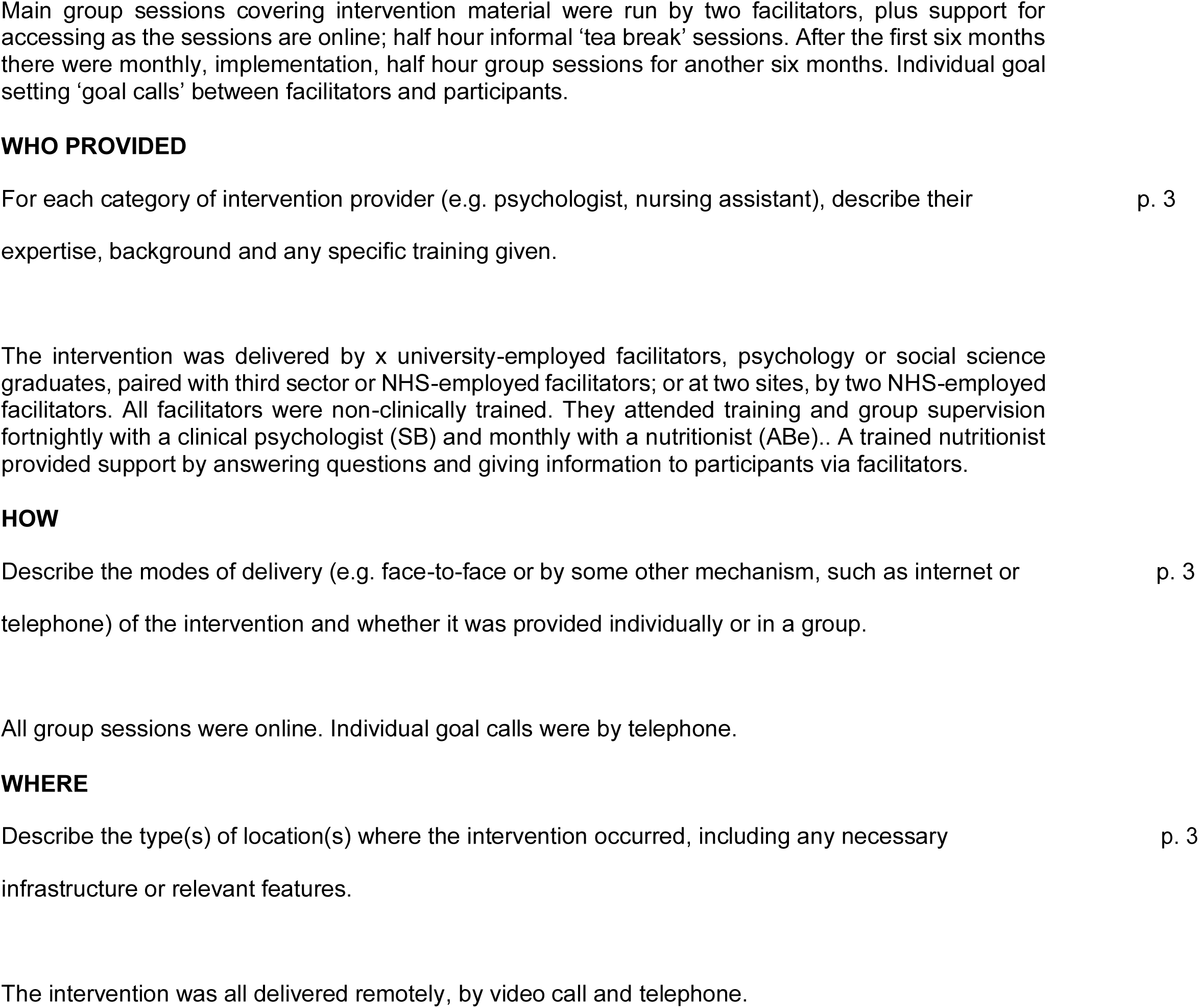
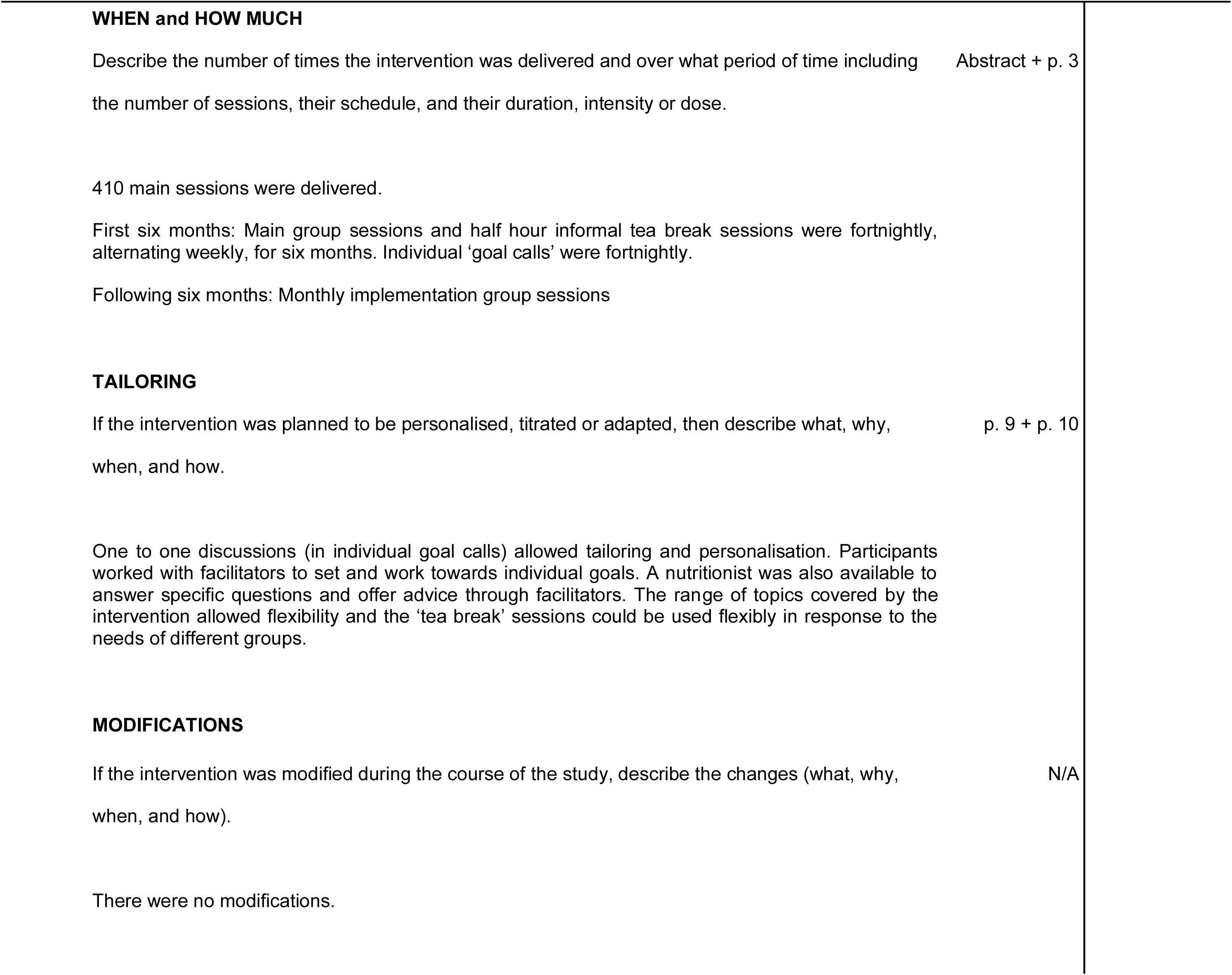
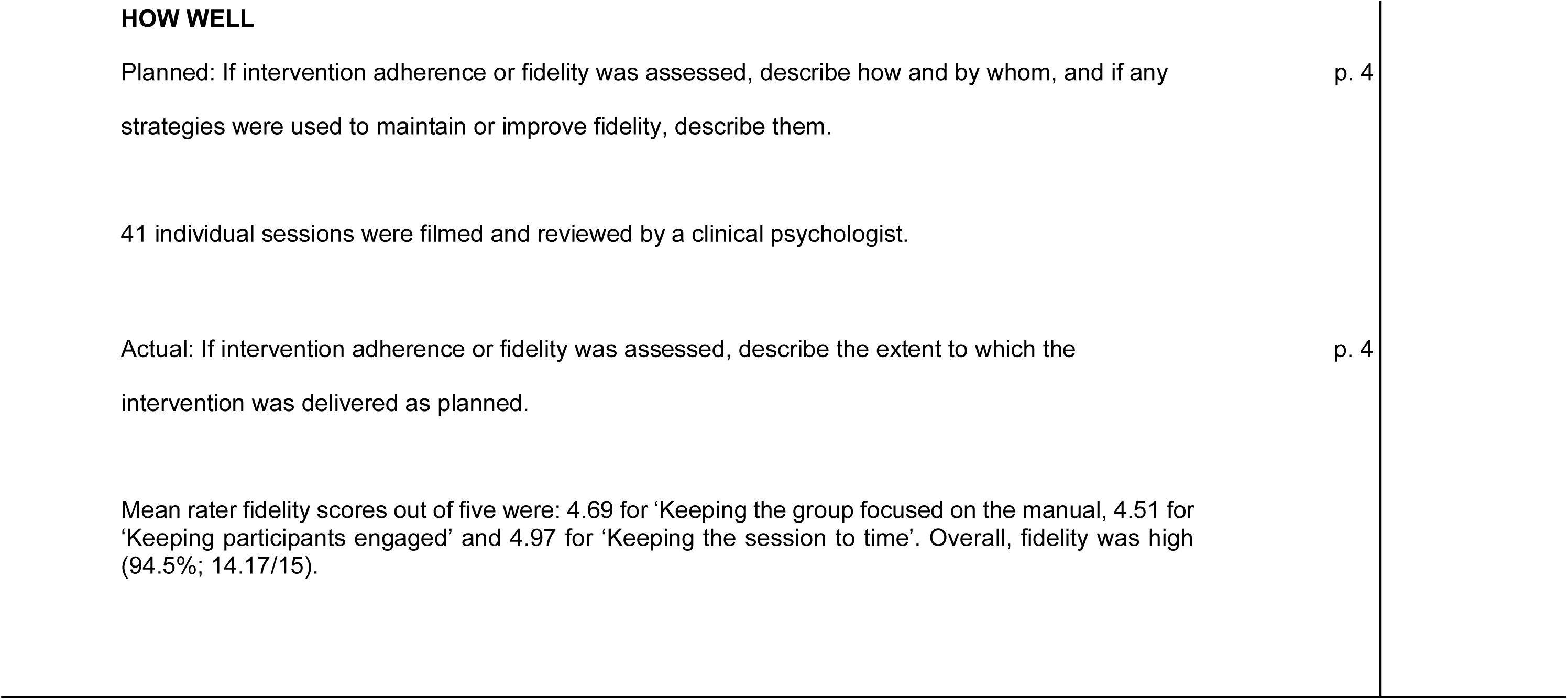

## Appendix B Participant interview topic guide

### APPLE-Tree study

#### Attendee interview Topic Guide

##### Introductions

Thank you for agreeing to take part in this interview. As you know I am a researcher from University College London and I am recording this interview.

##### Description of the research

You have been invited to attend the APPLE-Tree sessions over the past six months. I want to ask you about your experiences of it – the main sessions, the tea breaks, goal calls, catch-up sessions if you received any, and the cognitive training and app if you used them. We are interested in how APPLE-Tree helped you, or not, and for parts you did not use, what might have encouraged you to do so. We will use your suggestions to develop the programme for future participants. There are no right or wrong answers. We would particularly welcome any thoughts about things that could be done differently - please give us your honest opinion as this is what will help us most.

**Q. Please tell me about your experiences of the APPLE-Tree programme.**

**Prompts:**

○ *The initial set-up, being in the group, using zoom*
○ *The difference components: diet, exercise, planning new activities, wellbeing, looking after your mental and physical health*
○ *The tea breaks*
○ *Sending in photos and other material*
○ *Goal calls and setting goals*
○ *Plans to keep in touch with the group/ other participants after the main sessions have needed*
○ *The app*
○ *The cognitive training*

**Q. Were there things about the programme you found helpful?\*\***

**Prompts:**

○ *Can you tell me about that? Have you made any changes to your lifestyle or diet because of the groups? Do you think you will carry on with that change? What might make it easier/ harder to do so?*
○ *Anything else?*
○ *Do you think the group has helped your memory? (If so) Tell me what you have noticed. To what do you attribute that change?*

**Q. Was there anything about the programme you found unhelpful?\*\***

**Prompts:**

○ *Can you tell me about that?*
○ *Do you have any suggestions about how to improve the programme or how it could be different?*
○ *(for participants interviewed because they withdrew): can you tell me about your decision to leave the groups, what influenced your decision?*

**\*\*prompt if not discussed for group dynamics, how the group supported each other or not, eg was there a competitive element**

**Q. Before we finish, is there anything else you would like to add?**

## Appendix C APPLE-Tree logic model

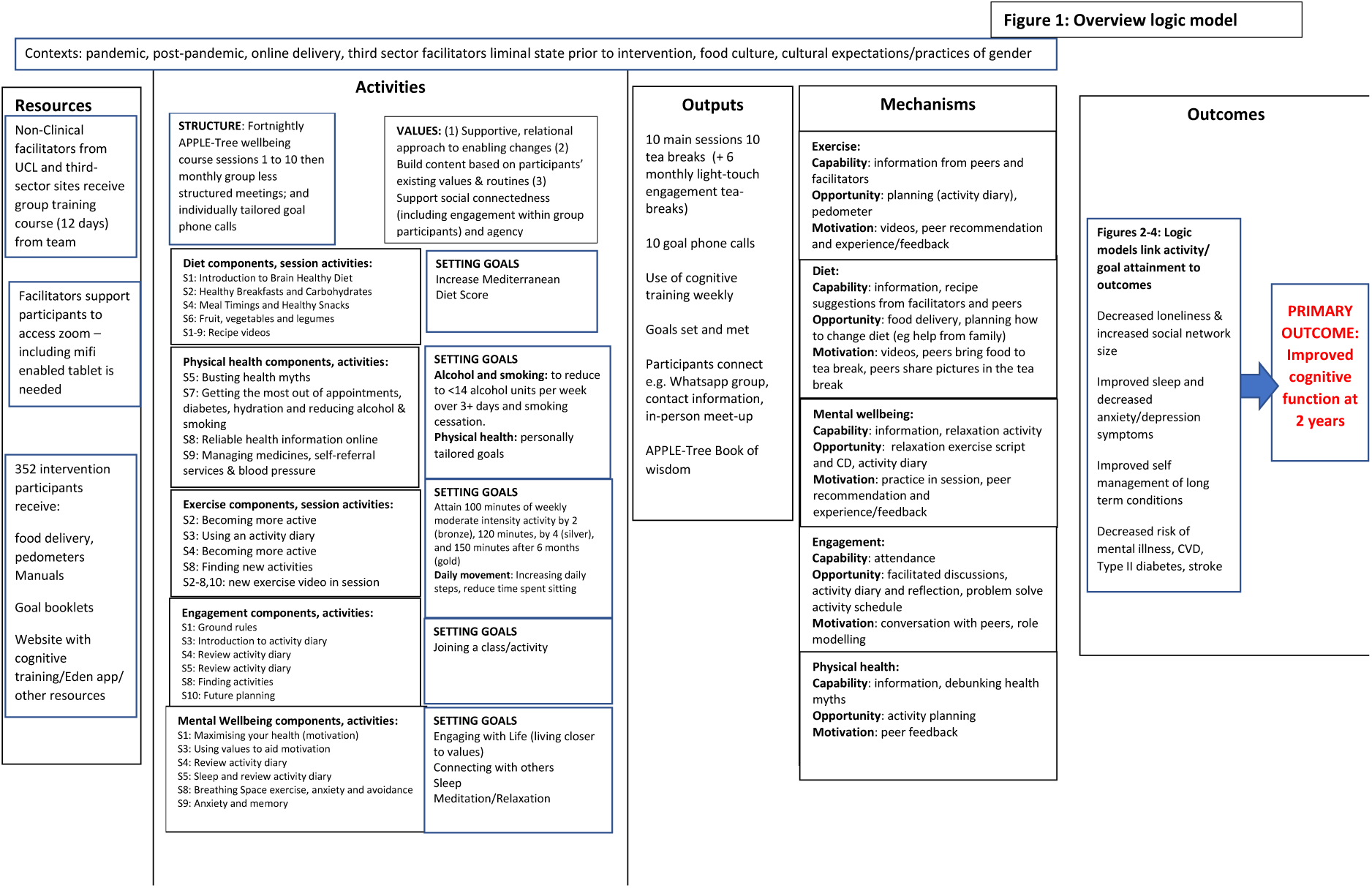

## Notes

### Competing Interest Statement

The authors have declared no competing interest.

### Clinical Trial

ISRCTN17325135

### Author Declarations

London (Camden and Kings Cross) Research Ethics Committee (Reference: 19/LO/0260) and UK Health Research Authority approved this study in April 2019. The protocol is registered (ISRCTN17325135).

